# Glucocorticoid-Induced Diabetes Mellitus in Tunisia: Incidence and Risk Factors in a Retrospective Cohort Study

**DOI:** 10.64898/2026.11.21.26368219

**Authors:** Youssef Bel Haj Hassen, Ibtissem Ben Nacef, Asma Kefi

**Author notes:** **Corresponding author:** Youssef Bel Haj Hassen, Faculty of Medicine of Tunis, University of Tunis El Manar, Tunis, Tunisia.

## Abstract

**Background:** Glucocorticoid-induced diabetes mellitus (GIDM) is a frequent but under-documented complication of systemic corticosteroid therapy. We aimed to estimate its incidence and identify patient- and treatment-related risk factors.

**Methods:** We conducted a retrospective, single-centre cohort study in the Department of Endocrinology and Internal Medicine A, Charles Nicolle Hospital, Tunis, between January 2000 and December 2020. Ninety-five adults without known diabetes (baseline HbA1c < 6.5%) starting corticosteroid therapy were followed for GIDM. Univariate predictors (p < 0.05) were entered into a multivariable model.

**Results:** Mean age was 42.6 years (M/F ratio 0.79). GIDM developed in 57 of 95 patients (60%), after a median of 20 days (range 7–120); 40% of cases occurred within the first month. The median initial dose was 60 mg/day prednisone-equivalent, and 84% received high-dose therapy. On univariate analysis, older age, family history of type 2 diabetes, higher baseline fasting glucose, higher body weight, and methylprednisolone boluses were associated with GIDM; sex, body mass index and hypertension were not. After adjustment, only family history of type 2 diabetes (RR 1.75, 95% CI 1.18–2.61) and higher baseline fasting glucose (RR 1.37 per 1 mmol/L, 95% CI 1.07–1.75) remained independent predictors.

**Conclusion:** GIDM occurred in three-fifths of patients, usually within the first month of therapy. Family history of type 2 diabetes and baseline fasting glucose independently identify patients warranting glycaemic surveillance from treatment initiation.

## Introduction

Glucocorticoids are among the most widely prescribed drugs in internal medicine, rheumatology, nephrology and transplantation, owing to their potent anti-inflammatory and immunosuppressive properties, but their long-term use carries substantial metabolic and cardiovascular toxicity [1,2]. Among these complications, drug-induced disturbance of glucose homeostasis is prominent, and glucocorticoids are among the most frequent pharmacological causes of diabetes [3].

By promoting hepatic gluconeogenesis, impairing insulin-mediated peripheral glucose uptake and inducing beta-cell dysfunction, glucocorticoids precipitate hyperglycaemia and, in a substantial proportion of treated patients, overt diabetes mellitus [4,5]. Glucocorticoid-induced diabetes mellitus (GIDM) is defined as new-onset hyperglycaemia meeting diagnostic criteria for diabetes in a patient receiving glucocorticoids and without previously known diabetes [6,7,8].

Reported frequencies of GIDM vary widely, from roughly 10% to more than 60%, the variation reflecting differences in the populations studied, the doses used, the diagnostic thresholds applied and, importantly, the intensity of glucose monitoring [7,9,10]. Despite this, GIDM remains under-recognised in routine practice, and data from North African populations are scarce [3,9].

We therefore conducted a retrospective cohort study in a Tunisian tertiary centre to estimate the incidence of GIDM among patients started on systemic corticosteroid therapy and to identify the patient- and treatment-related factors associated with its development.

## Patients and Methods

### Study design and population

This was a retrospective, single-centre cohort study carried out in the Department of Endocrinology and Internal Medicine A of Charles Nicolle Hospital Tunis. Records of patients started on systemic corticosteroid therapy between January 2000 and December 2020 were reviewed. Of 137 records identified, 42 were excluded because of incomplete or unusable files, leaving 95 patients for analysis.

Eligible patients were adults without previously known diabetes, with a baseline glycated haemoglobin (HbA1c) below 6.5% and a fasting plasma glucose below 7.0 mmol/L, who received systemic corticosteroid therapy (oral, with or without intravenous methylprednisolone) for at least 3 months at a dose of ≥ 15 mg/day prednisone-equivalent. Patients with pre-existing diabetes, those who had received corticosteroids in the preceding three months, and those with incomplete records were excluded.

### Definitions and data collection

GIDM was defined as new-onset diabetes in a patient without previously known diabetes, diagnosed during hospitalisation or follow-up by any of the following: a random plasma glucose ≥ 11.1 mmol/L, a fasting plasma glucose ≥ 7.0 mmol/L, or a glycated haemoglobin (HbA1c) ≥ 6.5% [8]. HbA1c, measured at baseline to exclude pre-existing diabetes, was also used during follow-up as one of the diagnostic criteria. For each patient we recorded demographic characteristics, family and personal medical history, the indication for corticosteroid therapy, the molecule, route, initial daily dose, cumulative dose and duration of treatment, the use and total dose of intravenous methylprednisolone boluses, and baseline anthropometric and metabolic parameters.

### Statistical analysis

Quantitative variables are summarised as means with standard deviations or medians with ranges, and qualitative variables as counts and percentages. Groups were compared using the Student t test or Mann–Whitney U test for continuous variables and the chi-square or Fisher exact test for categorical variables. Variables associated with GIDM at p < 0.05 in univariate analysis were entered into a multivariable model, and factors with p < 0.05 were retained as independently associated. Associations are reported as relative risks (RR) estimated by modified Poisson regression with robust error variance; relative risks for continuous variables are expressed per clinically meaningful increment. A two-sided p < 0.05 was considered significant.

## Results

### Population characteristics and indications

Ninety-five patients were included. The mean age was 42.6 years and the sex ratio (M/F) was 0.79. Baseline characteristics of patients who did and did not develop GIDM are shown in Table 1. The indications for corticosteroid therapy are shown in Table 2; systemic vasculitides and systemic autoimmune diseases together accounted for almost three-quarters of cases.

**Table 1.**
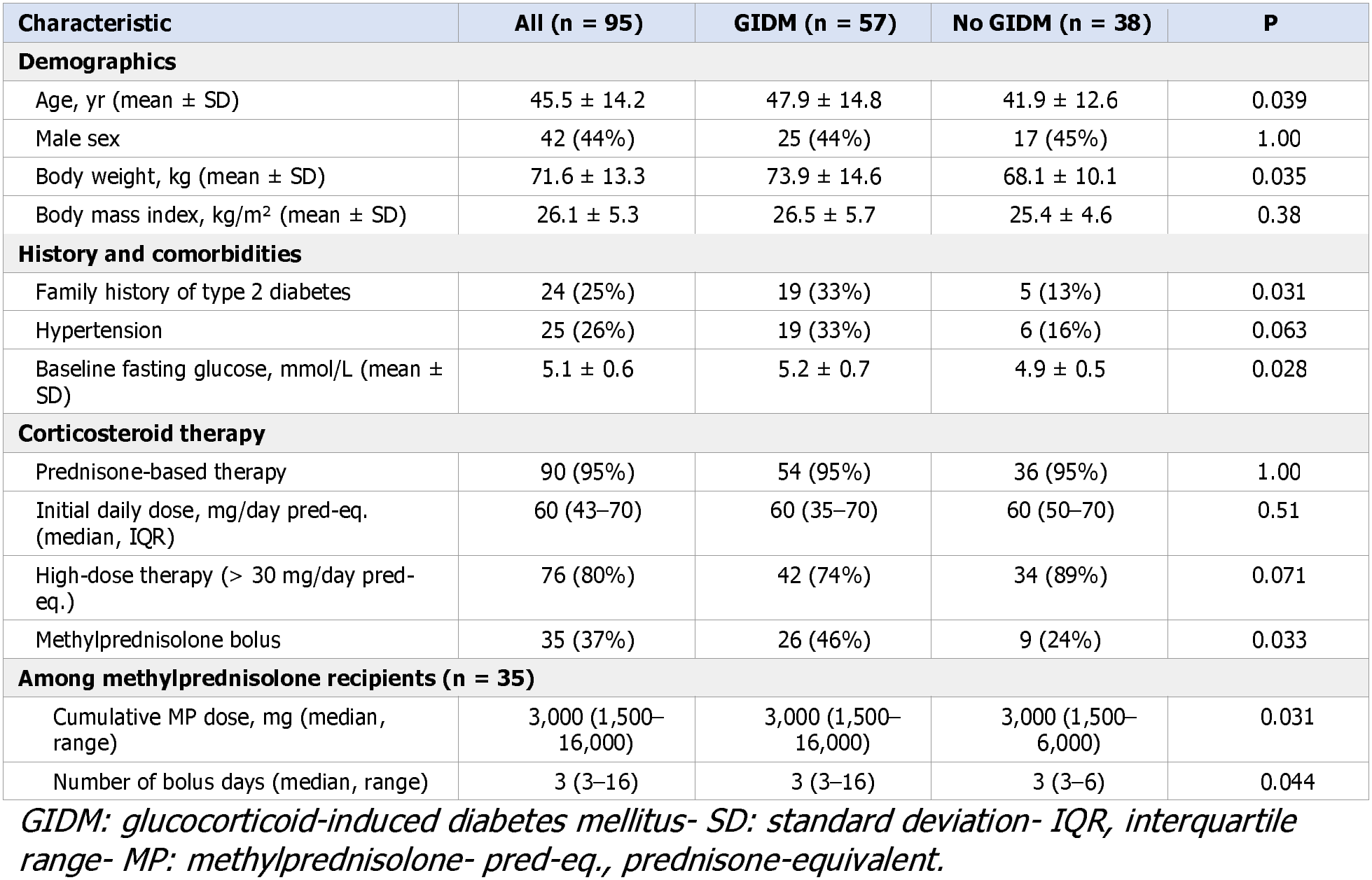
Baseline and corticosteroid-therapy characteristics, by GIDM status.

**Table 2.** Indications for corticosteroid therapy (n = 95).

| **Indication** | **n** | **%** |
|---|---|---|
| Systemic vasculitis | 39 | 41 |
| Systemic autoimmune disease | 31 | 33 |
| Organ-specific autoimmune disease | 4 | 4 |
| Granulomatosis | 3 | 3 |
| Autoimmune cytopenias | 5 | 5 |
| Other (incl. renal transplantation, n = 8) | 13 | 14 |
| **Total** | **95** | **100** |

### Corticosteroid therapy

Prednisone was the corticosteroid used in 90 patients (95%); the remainder received dexamethasone or hydrocortisone. The median initial dose was 60 mg/day prednisone-equivalent, and 76 patients (80%) received high-dose therapy, defined as more than 30 mg/day prednisone-equivalent. Intravenous methylprednisolone boluses were administered to 35 patients (37%). The median duration of corticosteroid therapy was 11 months. Methylprednisolone exposure was consistently associated with GIDM: bolus recipients developed GIDM more often than non-recipients (p = 0.033), and among the 35 recipients, both the cumulative methylprednisolone dose and the number of bolus days were higher in those who developed GIDM (p = 0.031 and p = 0.044, respectively). None of these remained independent after adjustment for patient-level factors.

### Incidence and characteristics of GIDM

GIDM developed in 57 of the 95 patients, giving a cumulative incidence of 60%. The median time to onset was 20 days (range 7–120), and 40% of cases occurred within the first 30 days of therapy. At diagnosis, the mean glucocorticoid dose was 50 mg/day prednisone-equivalent. Of 21 patients with baseline prediabetes, 16 (76%) progressed to GIDM. Most affected patients (94%) required pharmacological treatment in addition to lifestyle measures: oral agents in 45%, insulin in 33% and a combination in 16%; the remaining 6% were controlled with dietary measures alone.

### Factors associated with GIDM

On univariate analysis, older age, a family history of type 2 diabetes, a higher baseline fasting plasma glucose, a higher body weight and methylprednisolone bolus exposure were associated with GIDM (Table 3). Body mass index, sex, hypertension, the initial oral dose and the molecule used were not associated. In particular, body mass index did not differ significantly between patients who did and did not develop GIDM (26.5 vs 25.4 kg/m^2^, p = 0.38), and the association with hypertension did not reach significance (p = 0.063).

**Table 3.**
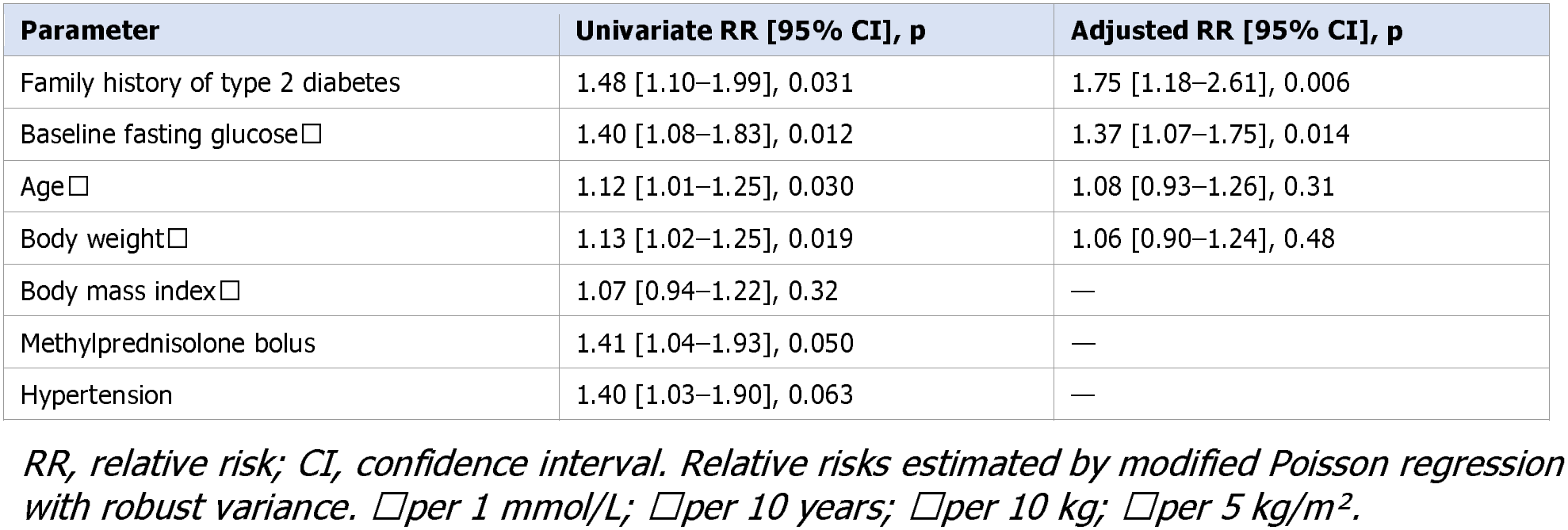
Factors associated with glucocorticoid-induced diabetes mellitus.

| **Parameter** | **Univariate RR [95% CI], p** | **Adjusted RR [95% CI], p** |
|---|---|---|
| Family history of type 2 diabetes | 1.48 [1.10–1.99], 0.031 | 1.75 [1.18–2.61], 0.006 |
| Baseline fasting glucose□ | 1.40 [1.08–1.83], 0.012 | 1.37 [1.07–1.75], 0.014 |
| Age□ | 1.12 [1.01–1.25], 0.030 | 1.08 [0.93–1.26], 0.31 |
| Body weight□ | 1.13 [1.02–1.25], 0.019 | 1.06 [0.90–1.24], 0.48 |
| Body mass index□ | 1.07 [0.94–1.22], 0.32 | — |
| Methylprednisolone bolus | 1.41 [1.04–1.93], 0.050 | — |
| Hypertension | 1.40 [1.03–1.90], 0.063 | — |

*RR, relative risk; CI, confidence interval. Relative risks estimated by modified Poisson regression with robust variance. □per 1 mmol/L; □per 10 years; □per 10 kg; □per 5 kg/m².*

All four variables significant at p < 0.05 in univariate analysis (family history, baseline glucose, age and body weight) were entered together into a single multivariable model (n = 70). Two remained independently associated with GIDM: a family history of type 2 diabetes (adjusted RR 1.75, 95% CI 1.18–2.61, p = 0.006) and a higher baseline fasting plasma glucose (adjusted RR 1.37 per 1 mmol/L, 95% CI 1.07–1.75, p = 0.014). Older age (adjusted RR 1.08 per 10 years, 95% CI 0.93–1.26, p = 0.31) and higher body weight (adjusted RR 1.06 per 10 kg, 95% CI 0.90–1.24, p = 0.48) did not remain independent after adjustment. Methylprednisolone bolus exposure was associated with GIDM on univariate analysis only.

## Discussion

In our Tunisian cohort, GIDM developed in 60% of patients started on systemic corticosteroids, most often within the first month of treatment. A family history of type 2 diabetes and a higher baseline fasting plasma glucose were the independent predictors; older age and higher body weight were associated on univariate analysis but not after adjustment. To our knowledge, this is among the first studies to characterise GIDM in a North African population [3].

Our incidence of 60% is high compared with most published studies, where reported rates range from about 10% to 65%. This wide range depends mainly on which patients are studied, the corticosteroid doses used, and how closely blood glucose is monitored, rather than on real biological differences [7,9,10]. Studies that test glucose systematically in high-dose, hospitalised patients report the highest rates: using repeated post-meal measurements, Katsuyama and colleagues found new diabetes in 65.6% of treated patients within four weeks [10]. In contrast, large studies based on primary-care records report much lower rates, because they count only patients coded as diabetic rather than testing everyone. In an English population study, systemic corticosteroids raised the risk of type 2 diabetes by about 85% (relative risk 1.85, 95% CI 1.74–1.97) [11]. Our patients were different: they were treated in hospital, mostly on high doses, and followed closely from the day corticosteroids were started. Our high rate therefore reflects a high-risk group that was watched carefully, not an overestimate.

The diabetes appeared early , after a median of 20 days, and in two-fifths of patients within the first month , which fits the way glucocorticoids affect blood sugar. They raise glucose within hours of a dose, typically causing an afternoon and after-meal peak when taken once each morning [4,12]. Since this effect is immediate [5,7], it makes sense that new diabetes appeared early in our high-dose patients rather than after months of treatment.

A family history of type 2 diabetes was the strongest predictor of GIDM. GIDM and type 2 diabetes share similar mechanisms, so an inherited tendency to one raises the risk of the other. Deutsch and colleagues found the same: in steroid-treated patients, a higher genetic risk score for type 2 diabetes predicted high blood sugar [15].

A higher baseline fasting glucose, even while still in the non-diabetic range, also predicted GIDM. Three-quarters of our patients with prediabetes at baseline developed diabetes. A higher baseline glucose has been shown to predict GIDM in patients with kidney disease [16] and in steroid-treated patients more generally [17], and checking it early helps catch diabetes sooner in the connective-tissue diseases that made up most of our cohort [18]. Because a fasting glucose is measured routinely, it offers a simple way to identify high-risk patients before treatment begins.

Older age was associated with GIDM on univariate analysis, consistent with the wider literature: age was a risk factor in the cohort of Katsuyama et al. [10], in population studies of steroid-related diabetes [11], in elderly patients specifically [13], and in the recent Oxford matched cohort [14]. Higher body weight was associated with GIDM on univariate analysis (heavier patients, 73.9 vs 68.1 kg), in keeping with studies that identify obesity and higher weight as risk factors for steroid-related diabetes [11,14] and with the threshold effect of obesity on insulin action [19]. As with age, however, weight did not remain independent after adjustment.

Treatment factors showed a similar pattern. Methylprednisolone boluses, and a higher total bolus dose, were linked to GIDM in univariate analysis, which fits the known dose-dependence of steroid diabetes [7,11,14]; in pemphigus, where very high doses are used, the risk of high blood sugar is about ten times higher [20]. Similar links between higher or longer exposure and GIDM have been reported in respiratory disease [21] and in lupus treated with high-dose steroids [22]. In our data, however, boluses were no longer linked to GIDM once family history and baseline glucose were taken into account.

A pattern worth highlighting is that every patient treated with corticosteroids for renal transplantation in our cohort developed GIDM. This is consistent with the prominent contribution of beta-cell dysfunction to post-transplantation diabetes [23] and with the elevated risk reported in primary renal disease [16], and it identifies transplant recipients as a group warranting particularly close glycaemic surveillance.

### Clinical implications

Our findings support a simple, pragmatic approach to risk stratification at the point of corticosteroid initiation. Patients who are older, who report a family history of type 2 diabetes, or whose baseline fasting glucose lies at the upper end of normal should be regarded as high-risk and offered glycaemic surveillance from the first days of therapy. Because glucocorticoid-induced hyperglycaemia is predominantly afternoon and post-prandial, fasting glucose alone may miss or delay the diagnosis. Post-prandial or afternoon capillary glucose monitoring, and where available continuous glucose monitoring, are more sensitive [4,12]. When pharmacological treatment is required, insulin remains the mainstay for moderate-to-severe glucocorticoid-induced hyperglycaemia, with regimens timed to the glucocorticoid’s glucose profile [6,24,25]. Structured early monitoring and treatment have been advocated across the settings in which glucocorticoids are used, from rheumatology to palliative care, yet clinical inertia frequently delays diagnosis [9,26,27].

### Strengths and limitations

This study has clear strengths: it addresses an under-studied North African population and follows patients without pre-existing diabetes from the start of treatment. It also has limitations. The modest sample size produced wide confidence intervals and limited the power to detect smaller effects. In addition, diabetes was diagnosed from random glucose and HbA1c that were not measured on a fixed schedule, so the timing of onset should be interpreted with caution.

### Future directions

Prospective, multicentre studies using protocolised post-prandial or continuous glucose monitoring are needed to establish the true incidence and timing of GIDM in this population and to clarify the contribution of adiposity. The incorporation of family history, and ultimately polygenic risk scores [15], into prospective risk-stratification tools, together with randomised evaluation of early monitoring and treatment strategies, would help translate these observations into preventive practice.

## Conclusion

Glucocorticoid-induced diabetes mellitus affected three-fifths of patients started on systemic corticosteroids in this Tunisian cohort, usually within the first month of treatment. A family history of type 2 diabetes and a higher baseline fasting glucose independently identified patients at risk, with older age and higher body weight as additional markers on univariate analysis. These readily available characteristics should prompt structured glycaemic monitoring from the initiation of corticosteroid therapy, enabling earlier detection and management of this common and clinically important complication.

## Data Availability

All data produced in the present study are available upon reasonable request to the authors

## Compliance with Ethical Standards

Ethics approval and consent to participate: The Ethics Committee of Charles Nicolle Hospital, Tunis, Tunisia, reviewed this study and waived the requirement for formal ethical approval and for individual informed consent, given its retrospective design and the use of anonymised data extracted from medical records. All procedures were conducted in accordance with the Declaration of Helsinki (1964) and its later amendments. This study did not involve animals.

## Author Contributions

All authors made equal contributions to the conception, design, data collection, analysis, interpretation, and manuscript preparation.

